## Supplemental Figures and Data for "Aerosol tracer testing in Boeing 767 and 777 aircraft to simulate exposure potential of infectious aerosol such as SARS-CoV-2"

### Aerosol tracer testing in the cabin of wide-bodied Boeing 767 and 777 aircraft to simulate exposure potential of infectious particulate such as SARS-CoV-2.

#### Supplemental Information and Figures

Sean M Kinahan[1,2][1]*, David B Silcott[1][3], Blake E Silcott[3], Ryan M Silcott[3], Peter J Silcott[3], Braden J Silcott[3], Steven L Distelhorst[2], Vicki L Herrera[1], Danielle N Rivera[2], Kevin K Crown[2], Gabriel A Lucero[2], Joshua L Santarpia[1,2]

[1] University of Nebraska Medical Center, 42nd and, Emile St, Omaha, NE 68198

[2] National Strategic Research Center, 6825 Pine Street, Omaha, NE 68106

[3] S3I LLC, 1135 Saffell Rd Reisterstown, MD 21136

[1] These authors contributed equally to this manuscript.

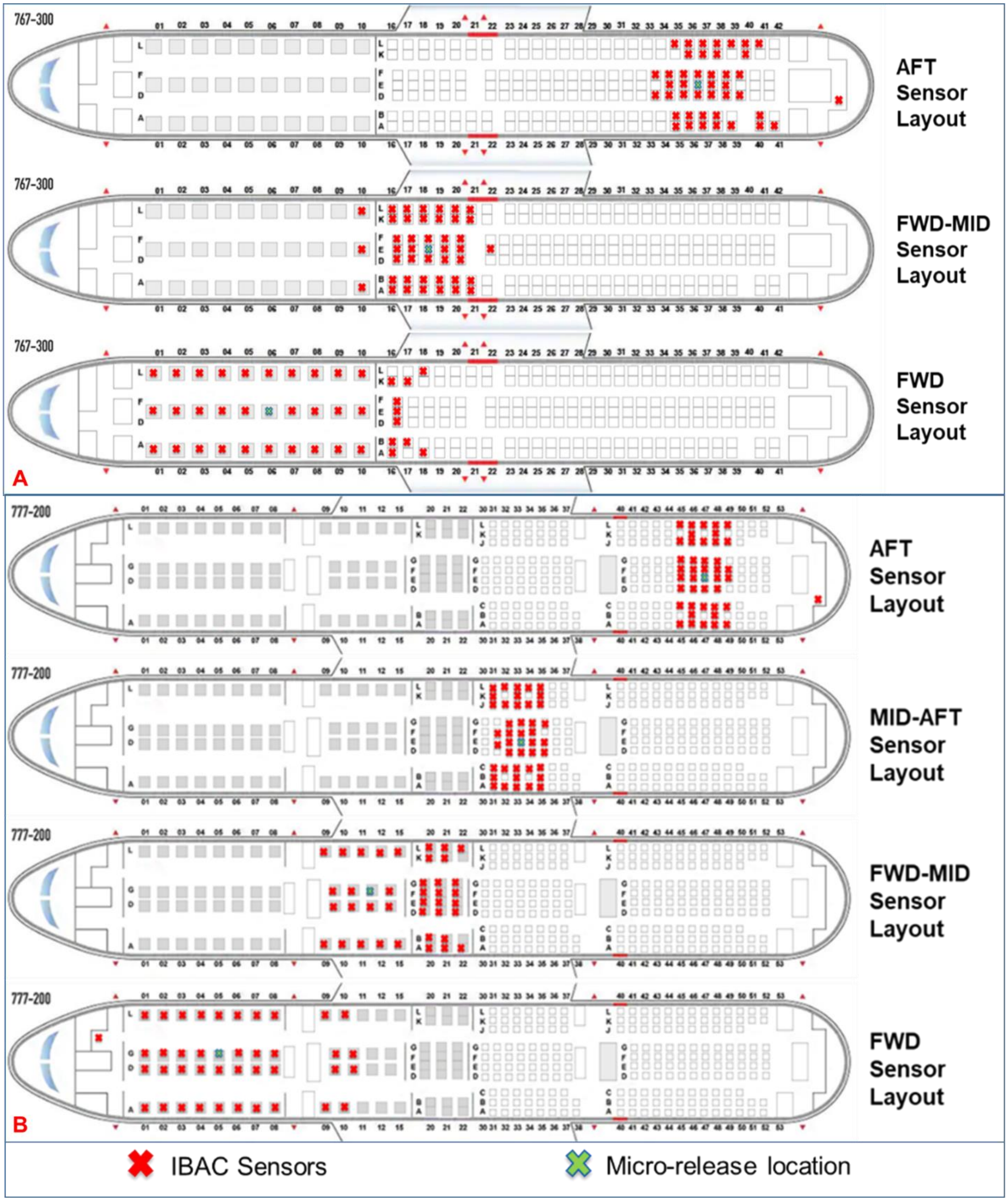


**Figure S1.** IBAC sensor layouts for each airframe and section tested. A) 767-300 sections and seats B) 777-200 sensors and seats. A single release seat is shown, but releases were done in multiple seats within a given row.

| 8/24/2020 | 777 Hangar Testing | | | |
|---|---|---|---|---|
| Inflight Tests | Airframe Section | Row/Seat Location | Gaspers | Mannequin Mask |
| Test 1 | AFT | 47A | OFF | OFF |
| Test 2 | AFT | 47A | OFF | OFF |
| Test 3 | AFT | 47B | OFF | OFF |
| Test 4 | AFT | 47B | OFF | OFF |
| Test 5 | AFT | 47C | OFF | OFF |
| Test 6 | AFT | 47C | OFF | OFF |
| Test 7 | AFT | 47D | OFF | OFF |
| Test 8 | AFT | 47D | OFF | OFF |
| Test 9 | AFT | 47E | OFF | OFF |
| Test 10 | AFT | 47E | OFF | OFF |
| Test 11 | AFT | 47F | OFF | OFF |
| Test 12 | AFT | 47F | OFF | OFF |
| Test 13 | AFT | 47G | OFF | OFF |
| Test 14 | AFT | 47G | OFF | OFF |
| Test 15 | AFT | 47J | OFF | OFF |
| Test 16 | AFT | 47J | OFF | OFF |
| Test 17 | AFT | 47K | OFF | OFF |
| Test 18 | AFT | 47K | OFF | OFF |
| Test 19 | AFT | 47L | OFF | OFF |
| Test 20 | AFT | 47L | OFF | OFF |
| Test 21 | FWD | 5A | OFF | OFF |
| Test 22 | FWD | 5D | OFF | OFF |
| Test 23 | FWD | 5G | OFF | OFF |
| Test 24 | FWD | 5L | OFF | OFF |
| Test 26 | FWD-MID | 11A | OFF | OFF |
| Test 27 | FWD-MID | 11D | OFF | OFF |
| Test 28 | FWD-MID | 11G | OFF | OFF |
| Test 29 | FWD-MID | 11L | OFF | OFF |
| Test 30 | MID-AFT | 33A | OFF | OFF |
| Test 31 | MID-AFT | 33B | OFF | OFF |
| Test 32 | MID-AFT | 33C | OFF | OFF |
| Test 33 | MID-AFT | 33D | OFF | OFF |
| Test 34 | MID-AFT | 33E | OFF | OFF |
| Test 35 | MID-AFT | 33F | OFF | OFF |
| Test 36 | MID-AFT | 33G | OFF | OFF |
| Test 37 | MID-AFT | 33J | OFF | OFF |
| Test 38 | MID-AFT | 33K | OFF | OFF |
| Test 39 | MID-AFT | 33L | OFF | OFF |

| 8/25/2020 | 777 Terminal Testing | | | | | |
|---|---|---|---|---|---|---|
| Inflight Tests | Airframe Section | Row/Seat Location | Conditions | Heat Blanket | Gaspers | Mannequin Mask |
| Test 1 | MID-AFT | 33E | Ground air on/Recirc off | ON | ON | OFF |
| Test 2 | MID-AFT | 33E | Ground air off / Recirc off | ON | OFF | OFF |
| Test 3 | MID-AFT | 33E | PACS on / Recirc on | ON | ON | OFF |
| Test 4 | MID-AFT | 33E | PACS on / Recirc on | OFF | ON | OFF |
| Test 5 | MID-AFT | 33E | PACS on / Recirc on | OFF | OFF | OFF |
| Test 6 | MID-AFT | 33E | PACS on / Recirc on | OFF | ON | ON |
| Test 7 | MID-AFT | 33E | PACS on / Recirc on | OFF | ON | ON |
| Test 8 | MID-AFT | 33E | PACS on / Recirc on | OFF | ON | OFF |
| Test 9 | FWD-MID | 11G | PACS on / Recirc on | OFF | OFF | OFF |
| Test 10 | FWD-MID | 11G | PACS on / Recirc on | OFF | ON | OFF |
| Test 11 | FWD-MID | 11G | PACS on / Recirc on | OFF | ON | OFF |
| Test 12 | FWD-MID | 11G | PACS on / Recirc on | OFF | ON | ON |
| Test 13 | FWD-MID | 11G | PACS on / Recirc on | OFF | ON | ON |
| Test 14 | FWD-MID | 11G | PACS on / Recirc on | OFF | ON | ON |
| Test 15 | FWD-MID | 11G | PACS on / Recirc on | OFF | ON | OFF |
| Test 16 | FWD-MID | 11G | PACS on / Recirc on | OFF | ON | OFF |
| Test 17 | FWD-MID | 11G | PACS on / Recirc on | OFF | ON | OFF |
| Test 18 | FWD-MID | 11G | PACS on / Recirc on | OFF | ON | OFF |
| Test 19 | FWD-MID | 11G | PACS on / Recirc on | OFF | ON | ON |
| Test 20 | FWD-MID | 11G | PACS on / Recirc on | OFF | ON | OFF |
| Test 21 | FWD-MID | 11G | PACS on / Recirc on | OFF | ON | ON |
| Test 22 | FWD-MID | 11G | PACS on / Recirc on | OFF | ON | OFF |
| Test 23 | FWD-MID | 11G | PACS on / Recirc on | OFF | ON | OFF |
| Test 24 | AFT | 47E | PACS on / Recirc on | OFF | OFF | OFF |
| Test 25 | AFT | 47E | PACS on / Recirc on | OFF | OFF | OFF |

| 8/26/2020 | 777 In-Flight Day 1 Testing | | | |
|---|---|---|---|---|
| Inflight Tests | Airframe Section | Row/Seat Location | Gaspers | Mannequin Mask |
| Test 1 | 47B | AFT | OFF | OFF |
| Test 2 | 47B | AFT | OFF | OFF |
| Test 3 | 47B | AFT | OFF | OFF |
| Test 4 | 47B | AFT | OFF | ON |
| Test 5 | 47B | AFT | OFF | ON |
| Test 6 | 47B | AFT | OFF | ON |
| Test 7 | 47E | AFT | OFF | OFF |
| Test 8 | 47E | AFT | OFF | OFF |
| Test 9 | 47E | AFT | OFF | OFF |
| Test 10 | 47E | AFT | OFF | ON |
| Test 11 | 47E | AFT | OFF | ON |
| Test 12 | 47E | AFT | OFF | ON |
| Test 13 | 47K | AFT | OFF | OFF |
| Test 14 | 47K | AFT | OFF | OFF |
| Test 15 | 47K | AFT | ON | OFF |
| Test 16 | 47K | AFT | ON | OFF |
| Test 17 | 47K | AFT | ON | OFF |
| Test 18 | 47K | AFT | ON | OFF |
| Test 19 | 33B | MID-AFT | OFF | OFF |
| Test 20 | 33B | MID-AFT | OFF | OFF |
| Test 21 | 33B | MID-AFT | OFF | OFF |
| Test 22 | 33B | MID-AFT | OFF | ON |
| Test 23 | 33B | MID-AFT | OFF | ON |
| Test 24 | 33B | MID-AFT | OFF | ON |
| Test 25 | 33E | MID-AFT | OFF | OFF |
| Test 26 | 33E | MID-AFT | OFF | OFF |
| Test 27 | 33E | MID-AFT | OFF | OFF |
| Test 28 | 33E | MID-AFT | OFF | ON |
| Test 29 | 33E | MID-AFT | OFF | ON |
| Test 30 | 33E | MID-AFT | OFF | ON |
| Test 31 | 33K | MID-AFT | OFF | OFF |
| Test 32 | 33K | MID-AFT | OFF | OFF |
| Test 33 | 33K | MID-AFT | OFF | OFF |

| 8/27/2020 | 777 In-Flight Day 2 Testing | | | |
|---|---|---|---|---|
| Inflight Tests | Airframe Section | Row/Seat Location | Gaspers | Mannequin Mask |
| Test 34 | FWD-MID | 11A | OFF | OFF |
| Test 35 | FWD-MID | 11A | OFF | OFF |
| Test 36 | FWD-MID | 11A | OFF | OFF |
| Test 37 | FWD-MID | 11A | OFF | ON |
| Test 38 | FWD-MID | 11A | OFF | ON |
| Test 39 | FWD-MID | 11A | OFF | ON |
| Test 40 | FWD-MID | 11G | OFF | OFF |
| Test 41 | FWD-MID | 11G | OFF | OFF |
| Test 42 | FWD-MID | 11G | OFF | OFF |
| Test 43 | FWD-MID | 11G | OFF | ON |
| Test 44 | FWD-MID | 11G | OFF | ON |
| Test 45 | FWD-MID | 11G | OFF | ON |
| Test 46 | FWD-MID | 11L | OFF | OFF |
| Test 47 | FWD-MID | 11L | OFF | OFF |
| Test 48 | FWD-MID | 11L | OFF | OFF |
| Test 49 | FWD-MID | 11L | OFF | ON |
| Test 50 | FWD-MID | 11L | OFF | ON |
| Test 51 | FWD-MID | 11L | OFF | ON |
| Test 52 | FWD | 5A | OFF | OFF |
| Test 53 | FWD | 5A | OFF | OFF |
| Test 54 | FWD | 5A | OFF | OFF |
| Test 55 | FWD | 5A | OFF | ON |
| Test 56 | FWD | 5A | OFF | ON |
| Test 57 | FWD | 5A | OFF | ON |
| Test 59 | FWD | 5G | OFF | OFF |
| Test 60 | FWD | 5G | OFF | OFF |
| Test 61 | FWD | 5G | OFF | OFF |
| Test 62 | FWD | 5L | OFF | OFF |
| Test 63 | FWD | 5L | OFF | OFF |
| Test 64 | FWD | 5L | OFF | OFF |

**Table S1-S4.** Boeing 777-200 Test conditions and timeline. All tests shown, data presented and analyzed here represents in-flight breathing tests.

| 8/28/2020 | 767 Hangar Testing | | | |
|---|---|---|---|---|
| Inflight Tests | Airframe Section | Row/Seat Location | Gaspers | Mannequin Mask |
| Test 1 | AFT | 37A | OFF | OFF |
| Test 2 | AFT | 37A | OFF | OFF |
| Test 3 | AFT | 37A | OFF | OFF |
| Test 4 | AFT | 37B | OFF | OFF |
| Test 5 | AFT | 37B | OFF | OFF |
| Test 6 | AFT | 37B | OFF | OFF |
| Test 7 | AFT | 37D | OFF | OFF |
| Test 8 | AFT | 37D | OFF | OFF |
| Test 9 | AFT | 37D | OFF | OFF |
| Test 10 | AFT | 37E | OFF | OFF |
| Test 11 | AFT | 37E | OFF | OFF |
| Test 12 | AFT | 37E | OFF | OFF |
| Test 13 | AFT | 37F | OFF | OFF |
| Test 14 | AFT | 37F | OFF | OFF |
| Test 15 | AFT | 37F | OFF | OFF |
| Test 16 | AFT | 37K | OFF | OFF |
| Test 17 | AFT | 37K | OFF | OFF |
| Test 18 | AFT | 37K | OFF | OFF |
| Test 19 | AFT | 37L | OFF | OFF |
| Test 20 | AFT | 37L | OFF | OFF |
| Test 21 | AFT | 37L | OFF | OFF |
| Test 22 | FWD | 5A | OFF | OFF |
| Test 23 | FWD | 5A | OFF | OFF |
| Test 24 | FWD | 5A | OFF | OFF |
| Test 26 | FWD | 7A | OFF | OFF |
| Test 27 | FWD | 7A | OFF | OFF |
| Test 28 | FWD | 6D | OFF | OFF |
| Test 29 | FWD | 6D | OFF | OFF |
| Test 30 | FWD | 6D | OFF | OFF |
| Test 31 | FWD | 5L | OFF | OFF |
| Test 32 | FWD | 5L | OFF | OFF |
| Test 33 | FWD | 5L | OFF | OFF |
| Test 34 | FWD-MID | 18A | OFF | OFF |
| Test 35 | FWD-MID | 18A | OFF | OFF |
| Test 36 | FWD-MID | 18A | OFF | OFF |
| Test 37 | FWD-MID | 18B | OFF | OFF |
| Test 38 | FWD-MID | 18B | OFF | OFF |
| Test 39 | FWD-MID | 18B | OFF | OFF |
| Test 40 | FWD-MID | 18D | OFF | OFF |
| Test 41 | FWD-MID | 18D | OFF | OFF |
| Test 42 | FWD-MID | 18D | OFF | OFF |
| Test 43 | FWD-MID | 18E | OFF | OFF |
| Test 44 | FWD-MID | 18E | OFF | OFF |
| Test 45 | FWD-MID | 18E | OFF | OFF |
| Test 46 | FWD-MID | 18F | OFF | OFF |
| Test 47 | FWD-MID | 18F | OFF | OFF |
| Test 48 | FWD-MID | 18F | OFF | OFF |
| Test 49 | FWD-MID | 18K | OFF | OFF |
| Test 50 | FWD-MID | 18K | OFF | OFF |
| Test 51 | FWD-MID | 18K | OFF | OFF |
| Test 52 | FWD-MID | 18L | OFF | OFF |
| Test 53 | FWD-MID | 18L | OFF | OFF |
| Test 54 | FWD-MID | 18L | OFF | OFF |

| 8/29/2020 | 767 Terminal Testing | | | | | |
|---|---|---|---|---|---|---|
| Inflight Tests | Airframe Section | Row/Seat Location | Cooling Conditions | Heat Blanket | Gaspers | Mannequin Mask |
| Test 1 | FWD-MID | 18E | Ground air ON/ Recirc ON | ON | ON | OFF |
| Test 2 | FWD-MID | 18E | Ground air ON/ Recirc ON | ON | ON | OFF |
| Test 3 | FWD-MID | 18E | Ground air ON/ Recirc ON | ON | ON | OFF |
| Test 4 | FWD-MID | 18E | Ground air ON/ Recirc ON | ON | OFF | OFF |
| Test 5 | FWD-MID | 18E | Ground air ON/ Recirc ON | ON | OFF | OFF |
| Test 6 | FWD-MID | 18E | Ground air ON/ Recirc ON | ON | OFF | OFF |
| Test 7 | FWD-MID | 18E | PACS ON / Recirc ON | ON | ON | OFF |
| Test 8 | FWD-MID | 18E | PACS ON / Recirc ON | ON | ON | OFF |
| Test 9 | FWD-MID | 18E | PACS ON / Recirc ON | ON | ON | OFF |
| Test 10 | FWD-MID | 18E | PACS ON / Recirc ON | OFF | ON | OFF |
| Test 11 | FWD-MID | 18E | PACS ON / Recirc ON | OFF | ON | OFF |
| Test 12 | FWD-MID | 18E | PACS ON / Recirc ON | OFF | ON | OFF |
| Test 13 | FWD-MID | 18E | PACS ON / Recirc ON | OFF | OFF | OFF |
| Test 14 | FWD-MID | 18E | PACS ON / Recirc ON | OFF | OFF | OFF |
| Test 15 | FWD-MID | 18E | PACS ON / Recirc ON | OFF | OFF | OFF |
| Test 16 | FWD-MID | 18E | PACS ON / Recirc ON | OFF | OFF | OFF |
| Test 17 | FWD-MID | 18E | PACS ON / Recirc ON | OFF | OFF | OFF |
| Test 18 | FWD-MID | 18E | PACS ON / Recirc ON | OFF | OFF | ON |
| Test 19 | FWD-MID | 18E | PACS ON / Recirc ON | OFF | OFF | ON |
| Test 20 | FWD-MID | 18E | PACS ON / Recirc ON | OFF | OFF | ON |
| Test 21 | FWD | 6D | PACS ON / Recirc ON | OFF | ON | OFF |
| Test 22 | FWD | 6D | PACS ON / Recirc ON | OFF | ON | OFF |
| Test 23 | FWD | 6D | PACS ON / Recirc ON | OFF | ON | OFF |
| Test 24 | FWD | 6D | PACS ON / Recirc ON | OFF | ON | OFF |
| Test 25 | FWD | 6D | PACS ON / Recirc ON | OFF | ON | ON |
| Test 26 | FWD | 6D | PACS ON / Recirc ON | OFF | ON | ON |
| Test 27 | FWD | 6D | PACS ON / Recirc ON | OFF | ON | ON |
| Test 28 | AFT | 37E | PACS ON / Recirc ON | OFF | ON | OFF |
| Test 29 | AFT | 37E | PACS ON / Recirc ON | OFF | ON | OFF |
| Test 30 | AFT | 37E | PACS ON / Recirc ON | OFF | ON | OFF |
| Test 31 | AFT | 37E | PACS ON / Recirc ON | OFF | ON | ON |
| Test 32 | AFT | 37E | PACS ON / Recirc ON | OFF | ON | ON |
| Test 33 | AFT | 37E | PACS ON / Recirc ON | OFF | ON | ON |

| 8/30/2020 | 767 In-Flight Day 1 Testing | | | | |
|---|---|---|---|---|---|
| Inflight Tests | Airframe Section | Row/Seat Location | Test Type | Gaspers | Mannequin Mask |
| Test 1 | AFT | 37B | Breathing | OFF | OFF |
| Test 2 | AFT | 37B | Breathing | OFF | OFF |
| Test 3 | AFT | 37B | Breathing | OFF | OFF |
| Test 4 | AFT | 37B | Breathing | OFF | ON |
| Test 5 | AFT | 37B | Breathing | OFF | ON |
| Test 6 | AFT | 37B | Breathing | OFF | ON |
| Test 7 | AFT | 37E | Breathing | OFF | OFF |
| Test 8 | AFT | 37E | Breathing | OFF | OFF |
| Test 9 | AFT | 37E | Breathing | OFF | OFF |
| Test 10 | AFT | 37E | Breathing | OFF | ON |
| Test 11 | AFT | 37E | Breathing | OFF | ON |
| Test 12 | AFT | 37E | Breathing | OFF | ON |
| Test 13 | AFT | 37E | Coughing | OFF | OFF |
| Test 14 | AFT | 37E | Coughing | OFF | OFF |
| Test 15 | AFT | 37E | Coughing | OFF | OFF |
| Test 16 | AFT | 37E | Coughing | OFF | ON |
| Test 17 | AFT | 37E | Coughing | OFF | ON |
| Test 18 | AFT | 37E | Coughing | OFF | ON |
| Test 19 | AFT | 37E | Coughing | OFF | OFF |
| Test 20 | AFT | 37K | Breathing | OFF | OFF |
| Test 21 | AFT | 37K | Breathing | OFF | OFF |
| Test 22 | AFT | 37K | Breathing | OFF | OFF |
| Test 23 | AFT | 37K | Breathing | OFF | ON |
| Test 24 | AFT | 37K | Breathing | OFF | ON |
| Test 25 | AFT | 37K | Breathing | OFF | ON |
| Test 26 | AFT | 37K | Coughing | OFF | OFF |
| Test 27 | AFT | 37K | Coughing | OFF | OFF |
| Test 28 | AFT | 37K | Coughing | OFF | OFF |
| Test 29 | AFT | 37K | Coughing | OFF | ON |
| Test 30 | AFT | 37K | Coughing | OFF | ON |
| Test 31 | AFT | 37K | Coughing | OFF | ON |
| Test 32 | FWD-MID | 18A | Breathing | OFF | OFF |
| Test 33 | FWD-MID | 18A | Breathing | OFF | OFF |
| Test 34 | FWD-MID | 18A | Breathing | OFF | OFF |
| Test 35 | FWD-MID | 18A | Breathing | OFF | ON |
| Test 36 | FWD-MID | 18A | Breathing | OFF | ON |
| Test 37 | FWD-MID | 18A | Breathing | OFF | ON |
| Test 38 | FWD-MID | 18A | Coughing | OFF | OFF |
| Test 39 | FWD-MID | 18A | Coughing | OFF | OFF |
| Test 40 | FWD-MID | 18A | Coughing | OFF | OFF |
| Test 41 | FWD-MID | 18A | Coughing | OFF | ON |
| Test 42 | FWD-MID | 18A | Coughing | OFF | ON |
| Test 43 | FWD-MID | 18A | Breathing | OFF | ON |
| Test 44 | FWD-MID | 18E | Breathing | OFF | OFF |
| Test 45 | FWD-MID | 18E | Breathing | OFF | OFF |
| Test 46 | FWD-MID | 18E | Breathing | OFF | OFF |
| Test 47 | FWD-MID | 18E | Breathing | OFF | ON |

| 8/31/2020 | 767 In-Flight Day 2 Testing | | | | |
|---|---|---|---|---|---|
| Inflight Tests | Airframe Section | Row/Seat Location | Test Type | Gaspers | Mannequin Mask |
| Test 48 | FWD-MID | 18E | Breathing | OFF | ON |
| Test 49 | FWD-MID | 18E | Breathing | OFF | ON |
| Test 50 | FWD-MID | 18L | Breathing | OFF | OFF |
| Test 51 | FWD-MID | 18L | Breathing | OFF | OFF |
| Test 52 | FWD-MID | 18L | Breathing | OFF | OFF |
| Test 53 | FWD-MID | 18L | Breathing | OFF | ON |
| Test 54 | FWD-MID | 18L | Breathing | OFF | ON |
| Test 55 | FWD-MID | 18L | Breathing | OFF | ON |
| Test 56 | FWD | 6A | Breathing | OFF | OFF |
| Test 57 | FWD | 6A | Breathing | OFF | OFF |
| Test 58 | FWD | 6A | Breathing | OFF | OFF |
| Test 59 | FWD | 6A | Breathing | OFF | ON |
| Test 60 | FWD | 6A | Breathing | OFF | ON |
| Test 61 | FWD | 6A | Breathing | OFF | ON |
| Test 62 | FWD | 6A | Coughing | OFF | OFF |
| Test 63 | FWD | 6A | Coughing | OFF | OFF |
| Test 64 | FWD | 6A | Coughing | OFF | OFF |
| Test 65 | FWD | 6A | Coughing | OFF | ON |
| Test 66 | FWD | 6A | Coughing | OFF | ON |
| Test 67 | FWD | 6A | Coughing | OFF | ON |
| Test 68 | FWD | 6D | Breathing | OFF | OFF |
| Test 69 | FWD | 6D | Breathing | OFF | OFF |
| Test 70 | FWD | 6D | Breathing | OFF | OFF |
| Test 71 | FWD | 6D | Breathing | OFF | ON |
| Test 72 | FWD | 6D | Breathing | OFF | ON |
| Test 73 | FWD | 6D | Breathing | OFF | ON |
| Test 74 | FWD | 6L | Breathing | OFF | OFF |
| Test 75 | FWD | 6L | Breathing | OFF | OFF |
| Test 76 | FWD | 6L | Breathing | OFF | OFF |
| Test 77 | FWD | 6L | Breathing | OFF | ON |
| Test 78 | FWD | 6L | Breathing | OFF | ON |
| Test 79 | FWD | 6L | Breathing | OFF | ON |
| Test 80 | FWD | 6L | Coughing | OFF | OFF |
| Test 81 | FWD | 6L | Coughing | OFF | ON |
| Test 82 | FWD | 6L | Coughing | OFF | OFF |
| Test 83 | FWD | 6L | Coughing | OFF | ON |
| Test 84 | FWD | 6L | Coughing | OFF | OFF |
| Test 85 | FWD | 6L | Coughing | OFF | ON |

**Table S5-S8.** Boeing 767-300 Test conditions and timeline. All tests shown, data presented and analyzed here represents in-flight breathing tests

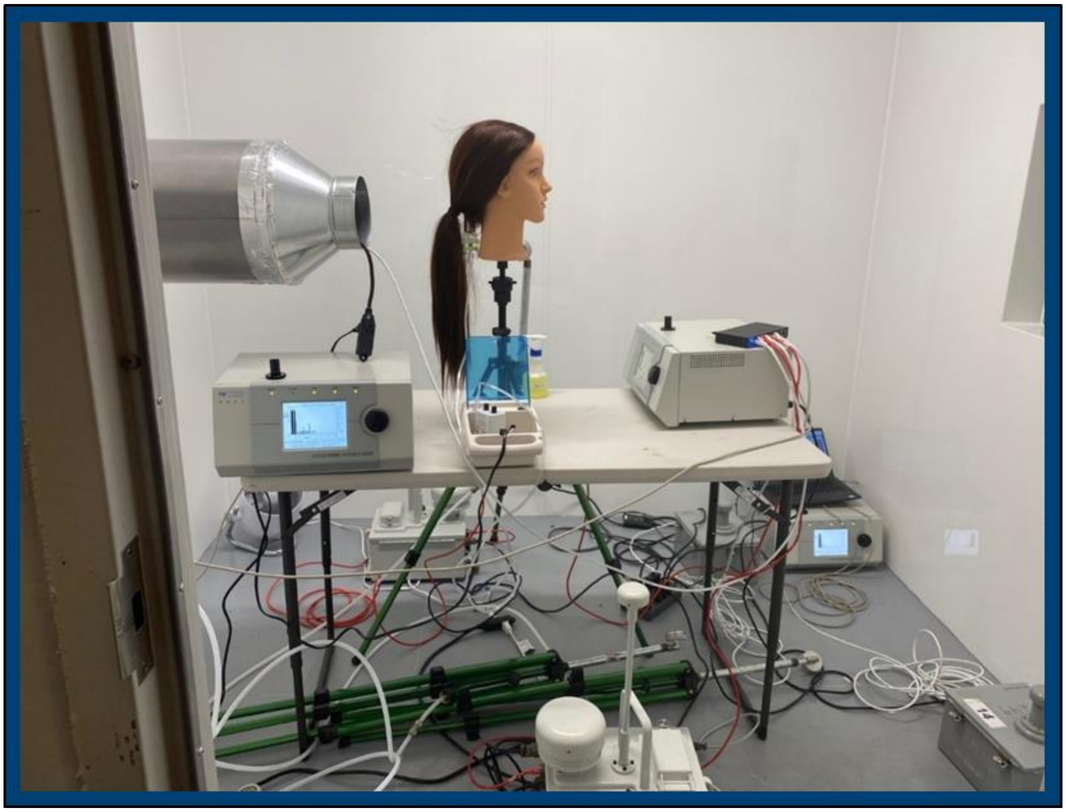

**Figure S2.** Chamber testing using a mannequin, three APS particle sizers, and four IBACs

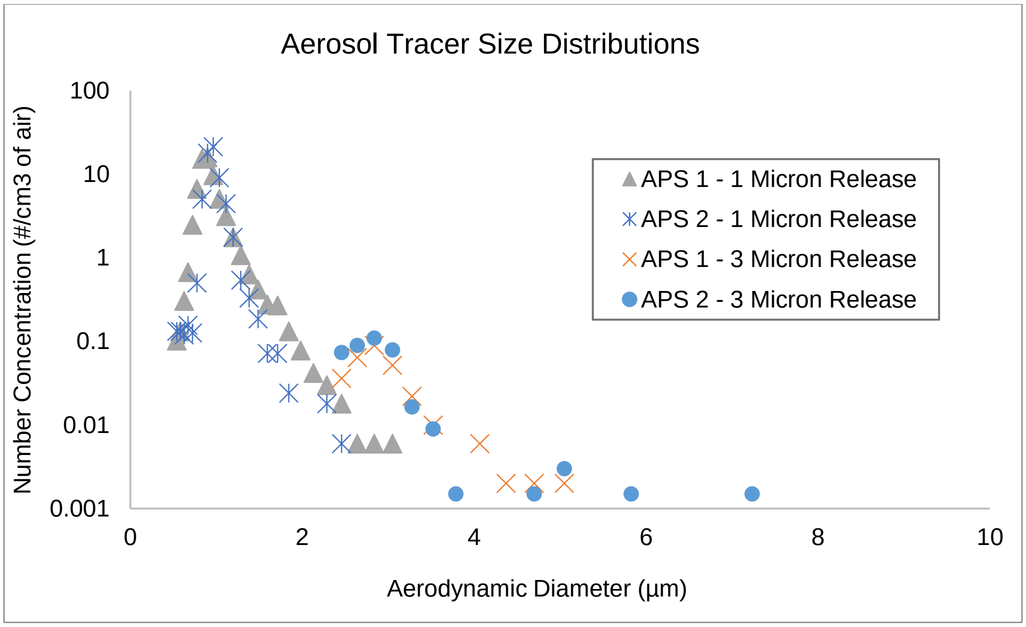


**Figure S3.** Characterization of Aerosol Tracer Particles at 1 and 3 µm.

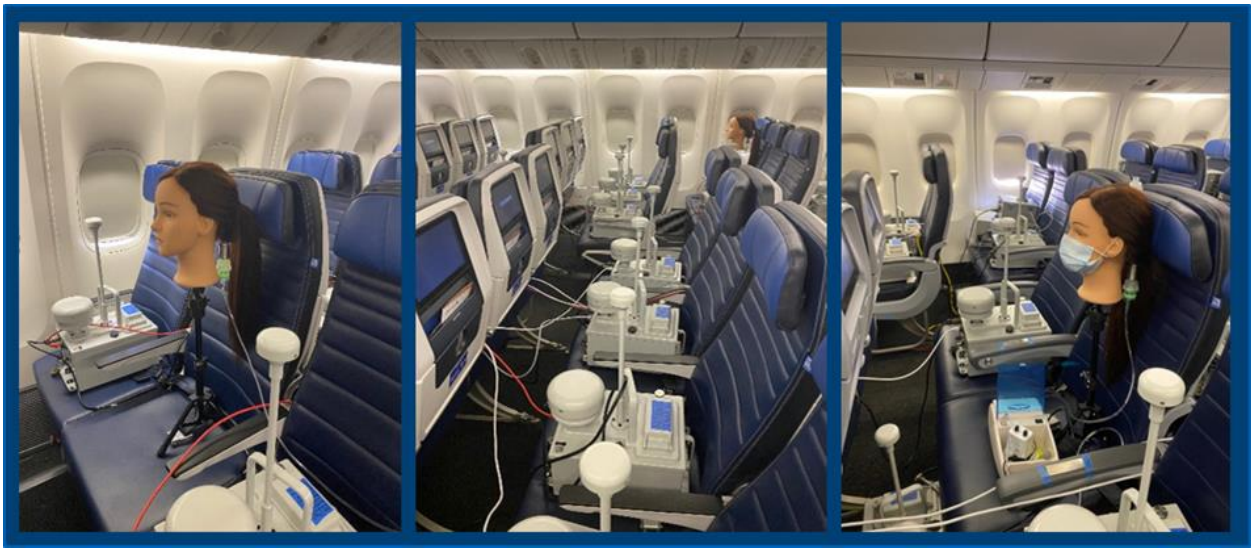

**Figure S4.** Visualization of mannequin and IBAC placement, with and without a mask.

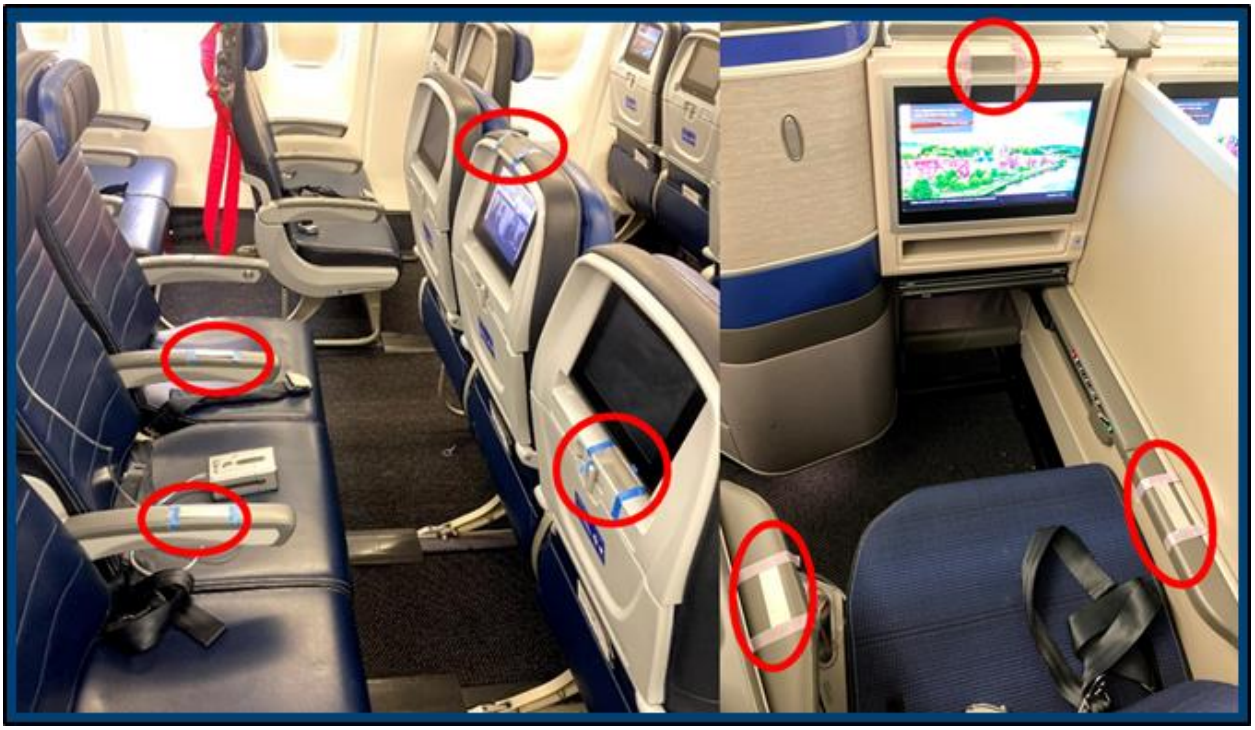

**Figure S5.** Example coupon locations highlighted in red. Left: Economy seat. Right: First class seat.

| Percent of Released Particles in 1 $ft^2$ (Surface Sample) or Integrated Collection at a Given Seat (Aerosol) | | | | | | | |
|---|---|---|---|---|---|---|---|
| Seat | Location | FWD | ±95% CI | MID-FWD | ±95% CI | MID-AFT | ±95% CI |
| 5D | Center Above IFE | 0.001% | 0.002% | 0.001% | 0.003% | 0.000% | 0.000% |
| 5D | Left Arm Rest | 0.001% | 0.001% | 0.001% | 0.002% | 0.000% | 0.000% |
| 5D | Right Arm Rest | 0.003% | 0.007% | 0.001% | 0.002% | 0.000% | 0.000% |
| 11D | Center Above IFE | 0.000% | 0.000% | 0.002% | 0.004% | 0.000% | 0.000% |
| 11D | Left Arm Rest | 0.000% | 0.000% | 0.001% | 0.003% | 0.000% | 0.001% |
| 11D | Right Arm Rest | 0.000% | 0.000% | 0.017% | 0.023% | 0.000% | 0.000% |
| 33D | Center Above IFE | 0.000% | 0.001% | 0.001% | 0.003% | 0.000% | 0.002% |
| 33E | Center Below IFE | 0.000% | 0.000% | 0.001% | 0.002% | 0.001% | 0.002% |
| 33E | Left Arm Rest | 0.000% | 0.001% | 0.002% | 0.004% | 0.018% | 0.060% |
| 33E | Right Arm Rest | 0.000% | 0.000% | 0.001% | 0.002% | 0.001% | 0.002% |
| 47E | Center Below IFE | 0.000% | 0.000% | 0.000% | #DIV/0! | 0.001% | 0.001% |
| 47E | Left Arm Rest | 0.000% | 0.000% | 0.000% | #DIV/0! | 0.000% | 0.000% |
| 47E | Right Arm Rest | 0.000% | 0.000% | 0.000% | #DIV/0! | 0.001% | 0.001% |
| 8D | Aerosol | 0.000% | 0.000% | 0.000% | 0.001% | 0.000% | 0.000% |
| 12D | Aerosol | 0.000% | 0.000% | 0.004% | 0.008% | 0.001% | 0.001% |
| 36E | Aerosol | 0.000% | 0.000% | 0.000% | 0.000% | 0.030% | 0.093% |
| 49D | Aerosol | 0.000% | 0.000% | 0.000% | 0.000% | 0.007% | 0.017% |
| Rear Galley | Aerosol | 0.000% | 0.001% | 0.000% | 0.000% | 0.002% | 0.006% |

**Table S9.** 777-200 DNA-Tagged Tracer Results (n=3), 95% CI based on standard error of the mean.

| Seat | Location | Percent of Released Particles in 1 Ft2 (Surface Sample) or Integrated Collection at a Given Seat (Aerosol) | | | | | |
|---|---|---|---|---|---|---|---|
| | | FWD | ±95% CI | MID | ±95% CI | AFT | ±95% CI |
| 6D | Left Arm Rest | 0.001% | 0.002% | 0.002% | 0.003% | 0.000% | 0.000% |
| 6D | Center Above IFE | 0.001% | 0.001% | 0.001% | 0.003% | 0.000% | 0.000% |
| 6D | Right Arm Rest | 0.003% | 0.009% | 0.003% | 0.008% | 0.002% | 0.008% |
| 6D | Marble Table | 0.003% | 0.004% | 0.005% | 0.005% | 0.000% | 0.001% |
| 18E | Left Arm Rest | 0.001% | 0.001% | 0.005% | 0.012% | 0.003% | 0.009% |
| 18E | Center Above IFE | 0.000% | 0.001% | 0.002% | 0.003% | 0.002% | 0.006% |
| 18E | Right Arm Rest | 0.000% | 0.001% | 0.001% | 0.003% | 0.000% | 0.001% |
| 18F | Center Below IFE | 0.000% | 0.000% | 0.001% | 0.002% | 0.000% | 0.002% |
| 26E | Tray Table | 0.000% | 0.001% | 0.002% | 0.003% | 0.003% | 0.008% |
| 37D | Center Above IFE | 0.000% | 0.001% | 0.000% | 0.000% | 0.001% | 0.003% |
| 37E | Left Arm Rest | 0.000% | 0.001% | 0.001% | 0.002% | 0.004% | 0.005% |
| 37E | Center Below IFE | 0.000% | 0.001% | 0.002% | 0.007% | 0.002% | 0.006% |
| 37E | Right Arm Rest | 0.000% | 0.001% | 0.001% | 0.002% | 0.001% | 0.001% |
| 5F | Aerosol | 0.004% | 0.012% | 0.000% | 0.000% | 0.000% | 0.000% |
| 22F | Aerosol | 0.000% | 0.000% | 0.000% | 0.001% | 0.000% | 0.001% |
| 31D | Aerosol | 0.000% | 0.000% | 0.004% | 0.008% | 0.001% | 0.004% |
| 40F | Aerosol | 0.000% | 0.000% | 0.000% | 0.000% | 0.012% | 0.016% |
| Rear Galley | Aerosol | 0.000% | 0.000% | 0.000% | 0.000% | 0.014% | 0.001% |

**Table S10.** 767-300 DNA-Tagged Tracer Results (n=3), 95% CI based on standard error of the mean

IBAC Raw Data Files and Additional Testing Breakdown Available At Figshare:

| Data Subset | FigShare DOI |
|---|---|
| USTRANSCOM 767 Inflight Master Spreadsheet with raw data. | https://doi.org/10.6084/m9.figshare.13537319.v1 |
| USTRANSCOM 777 Inflight Master Spreadsheet with raw data. | https://doi.org/10.6084/m9.figshare.13537349.v1 |
| USTRANSCOM 767 Hangar Master Spreadsheet with raw data. | https://doi.org/10.6084/m9.figshare.13537358.v1 |
| USTRANSCOM 777 Hangar Master Spreadsheet with raw data. | https://doi.org/10.6084/m9.figshare.13537379.v1 |
| USTRANSCOM 767 Terminal Master Spreadsheet with raw data. | https://doi.org/10.6084/m9.figshare.13537367.v1 |
| USTRANSCOM 777 Terminal Master Spreadsheet with raw data. | https://doi.org/10.6084/m9.figshare.13537385.v1 |
